# Long-read sequencing identifies *FGF14* repeat expansions in Parkinson’s disease

**DOI:** 10.1101/2025.08.14.25333596

**Authors:** Fulya Akçimen, Kensuke Daida, Lara M. Lange, Abraham Moller, Abigail Miano-Burkhardt, Laksh Malik, Kimberly Paquette, Pilar Alvarez Jerez, Jackson Mingle, Breeana Baker, Melissa Meredith, Cedric Kouam, Paige Jarreau, Androo Markham, Jessica Anderson, Miten Jain, Mark Chaisson, Mark Cookson, Bradford Casey, Hirotaka Iwaki, Sara Bandres-Ciga, Paula Saffie-Awad, Mike Nalls, Zih-Hua Fang, Andrew B. Singleton, Cornelis Blauwendraat, Kimberley J. Billingsley

## Abstract

Pathogenic GAA repeat expansions in *FGF14* are an established cause of late-onset cerebellar ataxia, but have not been linked to Parkinson’s disease (PD). Given emerging evidence that repeat expansions in ataxia-associated genes like *RFC1*, can contribute to atypical or familial forms of PD, we investigated whether *FGF14* expansions might play a similar role. Using long-read whole-genome sequencing on 411 individuals with PD and 197 neurologically healthy controls from the PPMI cohort, alongside 1,429 additional controls from the NIH CARD initiative, the 1000 Genomes Project, and the All of Us program, representing globally diverse populations. We identified pathogenic *FGF14* GAA repeat expansions in five individuals with PD and one control. All five individuals fit the clinical criteria of PD and showed typical patterns of neurodegeneration on DaTSCAN imaging; α-synuclein aggregation was confirmed by a positive seeding assay among four individuals with available data. These findings broaden the phenotypic spectrum of *FGF14* repeat-associated disease and suggest a rare, previously unrecognized genetic contributor to PD. To our knowledge, this is the first report implicating *FGF14* in PD and underscores the utility of long-read sequencing for detecting hidden forms of pathogenic variation in unresolved cases.

## Introduction

Most genetic studies of Parkinson’s disease (PD) focused on single-nucleotide variants (SNVs), while other forms of variation, such as short tandem repeats (STRs), are understudied, primarily due to technical limitations of short-read sequencing approaches^1^. Yet, pathogenic repeat expansions are established causes of several neurological disorders, including frontotemporal dementia (FTD), amyotrophic lateral sclerosis (ALS), and various forms of ataxia^2^. Interestingly, several repeat expansions primarily linked to other neurological diseases were also observed in PD patients, including those associated with spinocerebellar ataxia genes such as *ATXN2* and *ATXN3*^3,4^. In some PD genetic screening studies, repeat expansions were even observed at unexpectedly high frequencies. Further, biallelic AAGGG repeat expansions in *RFC1*, causing cerebellar ataxia, neuropathy, and vestibular areflexia syndrome^5^, have also been reported in a subset of individuals with PD in European cohorts^6,7^.

Recently, two independent studies identified pathogenic intronic GAA repeat expansions in *FGF14* in patients with late-onset ataxia. These expansions are considered fully penetrant when exceeding 300 repeats ((GAA) >300), and partially penetrant in the 250–300 repeat range^8,9^. While *FGF14* repeat expansions have been investigated in multiple systems atrophy - cerebellar type (MSA-C) cohorts, their potential role in PD has not yet been explored^10^,^11,12^.

In this study, we screened for pathogenic *FGF14* (GAA) repeat expansions in individuals with PD by leveraging Oxford Nanopore Technologies (ONT) long-read whole-genome sequencing (WGS) data. Our analysis included 411 PD cases from the Parkinson’s Progression Markers Initiative (PPMI) and 1,626 controls obtained from PPMI, NIH’s CARD Long-read initiative, All of Us, and 1000 Genomes Project participants. Additionally, we aimed to characterize repeat motifs in identified carriers, including alleles within the reduced penetrance range (250–300 repeats), and characterize alternative non-pathogenic repeat configurations, including (GAAGGA), (GAACGA), and composite motifs such as (GAA) (CGA).

## Methods

### Cohort information

We obtained frozen blood samples from PPMI (https://www.ppmi-info.org/) and accessed existing long-read WGS from the North American Brain Expression Consortium (NABEC), the Human Brain Collection Core (HBCC), All of Us Research Program release 8 (https://www.researchallofus.org/), and the 1000 Genomes Project (https://ftp.1000genomes.ebi.ac.uk/vol1/ftp/data_collections/1KG_ONT_VIENNA/) datasets. In total, the data comprised 411 PD cases and 1,626 neurologically healthy controls. All PD cases were diagnosed according to the UK PD Society Brain Bank^13^ criteria. Demographic characteristics of the other cohorts are shown in Supplementary Table 1.

### Long-read sequencing-based screening of *FGF14* repeat expansion

We screened three reference cohorts: (1) 204 European-ancestry controls from the NABEC and 133 African-ancestry controls from HBCC, both generated as part of the CARD Long-Read Initiative; (2) 908 globally diverse individuals from the 1000 Genomes Long-Read Project; and (3) 184 European-ancestry healthy controls from the All of Us long-read sequencing dataset.

Sequencing, quality control, alignment, variant calling and methylation analysis for the PPMI dataset are described in detail in the Supplementary Methods. Sequencing and downstream processing for the NIH’s CARD Long-read initiative cohorts, specifically the HBCC and NABEC samples, were previously described elsewhere^14^. In addition, we utilized long-read WGS data from the All of Us Research Program^15^ and 1000 Genomes Project ONT Panel^15,16^.

The length of the *FGF14* GAA repeat expansion was estimated using *Straglr*, a tool that enables both repeat sizing and the detection of alternative motifs within a defined region^17^. The repeat locus was defined as chr13:102161576-102161726 (hg38). For each sample, the (GAA) repeat length was calculated as the average of the repeat lengths from the ten reads with the longest observed expansions. For All of Us samples, where allelic depth was lower, estimates were based on the five longest reads. To assess the presence of interruptions or alternative repeat motifs, we generated waterfall plots using RepeatAnalysis tools (https://github.com/PacificBiosciences/apps-scripts/tree/master/RepeatAnalysisTools).

## Results

### Identifying carriers of the *FGF14* GAA repeat expansions

We screened for intronic GAA repeat expansions in *FGF14* using ONT long-read WGS in individuals from the PPMI dataset. Sequencing quality metrics, including read N50, median coverage, and total data yield, are summarized in Supplementary Table 2. An overview of the study design is presented in Figure 1.

**Figure 1.**
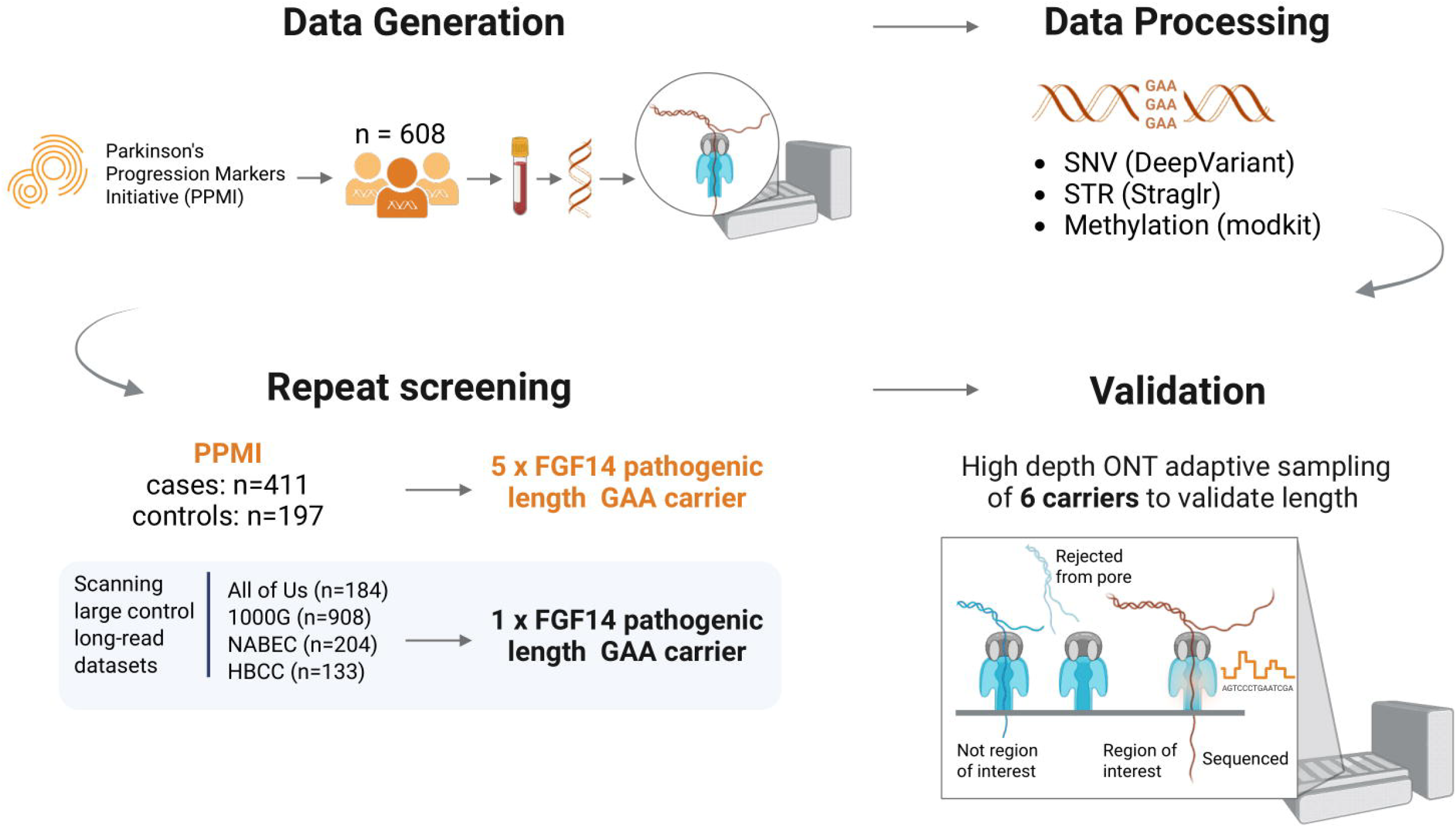
Study design for *FGF14* repeat expansion screening using long-read sequencing. Schematic overview of the analytical workflow and study rationale, highlighting the use of ONT sequencing to identify pathogenic GAA repeat expansions in *FGF14*. Created with BioRender.com

In the PPMI cohort, we identified five PD patients of European ancestry carrying *FGF14* GAA expansions above the pathogenic threshold of 300 GAA repeats (Figure 2), while no carriers were identified in the PPMI controls. These expansions were not accompanied by known pathogenic SNVs in established genes linked to monogenic PD (including *GBA1, LRRK2, SNCA, DJ-1, FBXO7, PINK1, PRKN, PLA2G6, VPS13C*, and *VPS35*). Only one individual (Patient 1) carried a variant in the PD risk gene *GBA1* (p.Leu483Pro). Among the control reference datasets, we identified one individual of European ancestry from the NABEC control brain dataset carrying an *FGF14* GAA expansion longer than 300 GAA repeats, exceeding the pathogenic threshold. This was a male individual who died in his 80s without any reported neurological symptoms. All six individuals, including the five PPMI PD patients and the NABEC control, were further validated by adaptive sampling long-read sequencing. The average depth of coverage over the *FGF14* repeat region, based on adaptive sampling long-read sequencing, was 55×, 125×, 90×, 97×, and 117× for PPMI patients 1 through 5, respectively, and 114× in the NABEC control (Figure 2).

**Figure 2.**
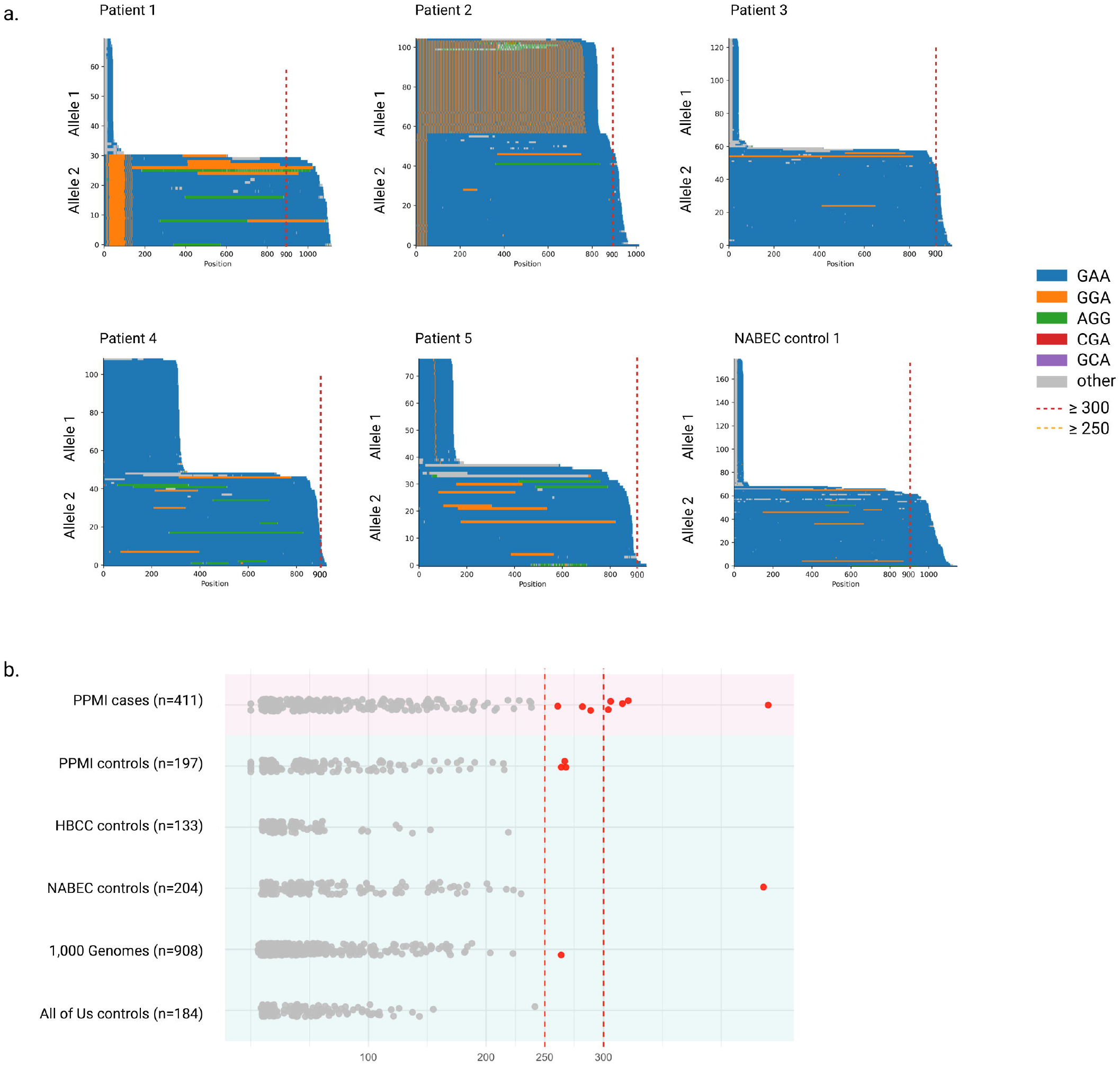
Detection of Pathogenic *FGF14* (GAA)n repeat expansions in Parkinson’s disease cases. **a)** Waterfall plots displaying the repeat lengths observed in five PD cases carrying fully penetrant (GAA) repeat expansion ≥300 repeat units, as determined by adaptive sampling long-read sequencing. **b)** Swimlane plot showing the distribution of *FGF14* (GAA) repeat lengths in the PPMI cases (n=411), the PPMI controls (n=197), in the NABEC/HBCC control cohorts (n=317) comprising individuals of European and African and African-admixed ancestry, the 1000 Genomes Project control cohort (n=908) comprising individuals of mixed ancestry, and the All of Us biobank participants (n=184) comprising individuals of European ancestry. Dashed red vertical lines denote the thresholds for reduced penetrance (250 repeat units) and full penetrance (300 repeat units), respectively.

A summary of *FGF14* repeat lengths, including the presence of alternative motifs in the repeat region, is provided for a total of 411 PD cases and 1,626 controls across all investigated cohorts in Supplementary Table 2. Our analysis in a subset of 386 PD cases and 722 controls of European ancestry show that carrying uninterrupted *FGF14* GAA expansions of ≥300 repeats is associated with PD (Fisher’s exact test; p = 0.022), while having ≥250 repeats is not associated with disease risk (Fisher’s exact test; p = 0.31). Repeat length modeled as a continuous variable was also nominally associated with PD risk (OR = 1.003, 95% CI: 1.000–1.005, p = 0.015).

We further investigated a potential correlation between repeat length and age at onset, as reported by previous studies in ataxia cohorts. Among the five identified *FGF14* expansion carriers, ages at onset ranged from 30s to 60s years. There was no correlation between repeat length and age at onset (R^2^ = 0.02, P = 0.85, Supplementary Figure 3). In addition, to examine whether increased repeat length is associated with earlier age at onset among patients, we performed a linear regression analysis. No significant associations were found between repeat length and age at onset (R^2^ = 0.01, P = 0.06) (Supplementary Figure 3).

To better interpret these findings, we examined *FGF14* repeat length variation across several large long-read control datasets. In these datasets, uninterrupted *FGF14* GAA repeat lengths ranged from 9 to 436 in NABEC, 10 to 219 in HBCC, 9 to 242 in All of Us, and 6 to 264 in 1000 Genomes samples (Figure 2, Supplementary Table 2). These results define the normal range of *FGF14* repeat length across ancestrally diverse control populations and provide a critical reference framework for interpreting expansions in patient cohorts. Collectively, our findings reinforce the pathogenic size threshold of ≥300 repeats and establish a foundation for future studies investigating the role of *FGF14* in neurodegenerative diseases.

*FGF14* locus was hypermethylated in blood across all samples from PPMI, while brain-derived DNA from the cerebellum of the NABEC carrier showed hypomethylation without allele-specific differences (Supplementary Figure 4).

### Characterization of *FGF14* repeat motif structure in Parkinson’s disease

We identified seven individuals, three PD patients and four controls (three from the PPMI cohort and one from the 1000 Genomes Project), carrying *FGF14* (GAA) repeat expansions in the reduced penetrance range of 250–300 repeats. In addition to pure GAA expansions, both Rafehi et al. and Pellerin et al. reported complex or interrupted motifs^8,9^, such as composite structures involving (GAAGGA) or [(GAA)4(GCA)1], raising the possibility that sequence composition, not just repeat length, may influence pathogenicity^18^. To investigate whether similar motif variability is present in PD, we characterized the repeat structure at the *FGF14* locus in ONT long-read WGS data from fully penetrant and reduced penetrance expansion carriers in cases and control cohorts. We identified 11 controls carrying the (GAAGGA) motif, two from the PPMI cohort, four from NABEC, three from the 1000 Genomes Project (including one European [Utah residents with Northern and Western European ancestry], one American [Puerto Rican in Puerto Rico], and one South Asian ancestry [Sri Lankan Tamil in the UK]), and two from the All of Us European ancestry cohort. Additionally, we identified one PPMI case and 13 controls from the 1000 Genomes Project carrying the (GAAGCA) motif, six of East Asian and seven of American ancestry, as well as a composite (GAA) (GCA) motif in a 1000 Genomes participant of East Asian ancestry.

### Clinical features of the Parkinson’s disease patient carrying the pathogenic *FGF14 GAA expansion*

The clinical features of all identified carriers are summarized in Table 1 and will be reported in detail below.

**Table 1.**
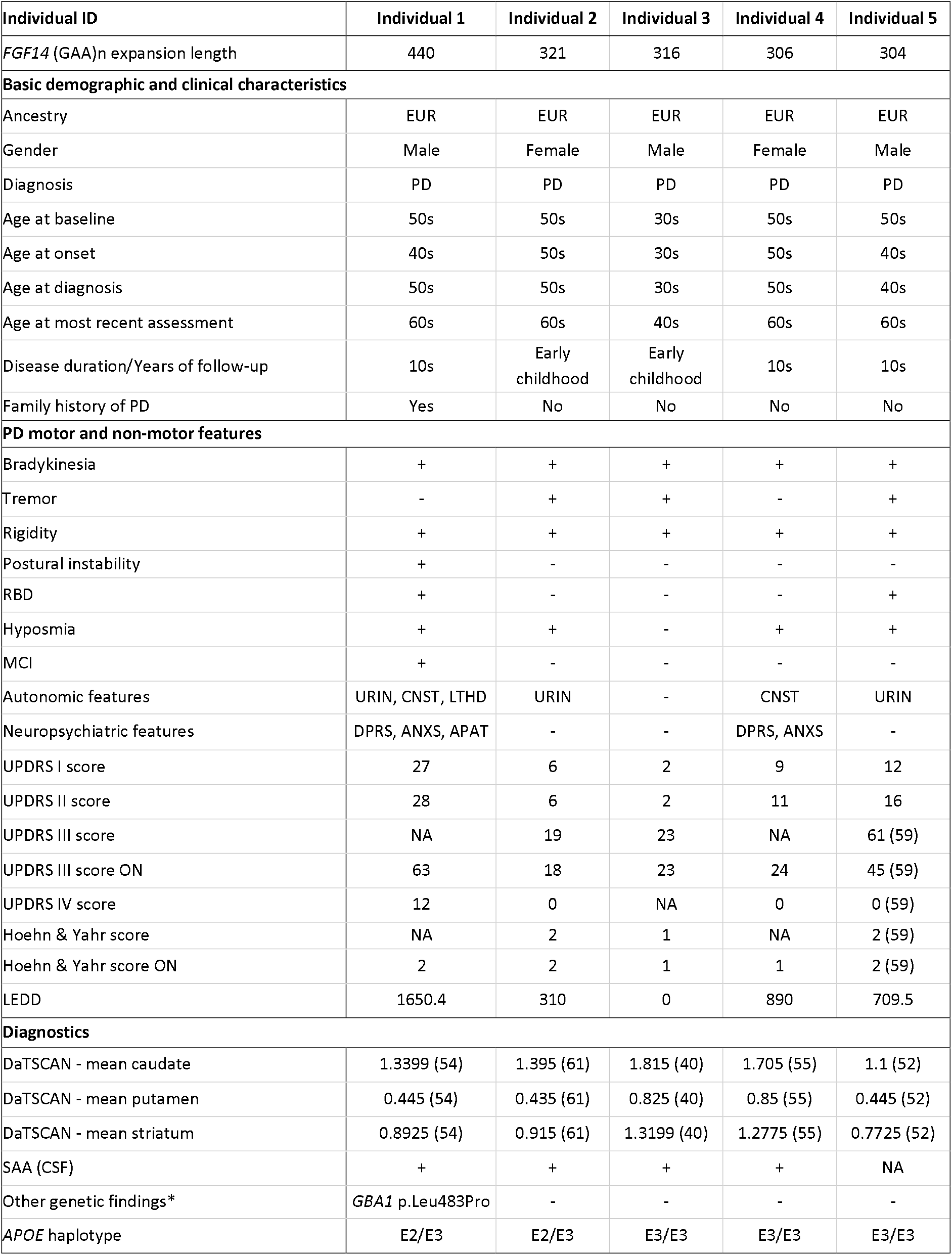
Clinical characteristics of the PD patients carrying a pathogenic length FGF14 GAA expansion.

| Individual ID | Individual 1 | Individual 2 | Individual 3 | Individual 4 | Individual 5 |
| --- | --- | --- | --- | --- | --- |
| *FGF14* (GAA)<sub>n</sub> expansion length | 440 | 321 | 316 | 306 | 304 |
| **Basic demographic and clinical characteristics** |  |  |  |  |  |
| Ancestry | EUR | EUR | EUR | EUR | EUR |
| Gender | Male | Female | Male | Female | Male |
| Diagnosis | PD | PD | PD | PD | PD |
| Age at baseline | 50s | 50s | 30s | 50s | 50s |
| Age at onset | 40s | 50s | 30s | 50s | 40s |
| Age at diagnosis | 50s | 50s | 30s | 50s | 40s |
| Age at most recent assessment | 60s | 60s | 40s | 60s | 60s |
| Disease duration/Years of follow-up | 10s | Early childhood | Early childhood | 10s | 10s |
| Family history of PD | Yes | No | No | No | No |
| **PD motor and non-motor features** |  |  |  |  |  |
| Bradykinesia | + | + | + | + | + |
| Tremor | - | + | + | - | + |
| Rigidity | + | + | + | + | + |
| Postural instability | + | - | - | - | - |
| RBD | + | - | - | - | + |
| Hyposmia | + | + | - | + | + |
| MCI | + | - | - | - | - |
| Autonomic features | URIN, CNST, LTHD | URIN | - | CNST | URIN |
| Neuropsychiatric features | DPRS, ANXS, APAT | - | - | DPRS, ANXS | - |
| UPDRS I score | 27 | 6 | 2 | 9 | 12 |
| UPDRS II score | 28 | 6 | 2 | 11 | 16 |
| UPDRS III score | NA | 19 | 23 | NA | 61 (59) |
| UPDRS III score ON | 63 | 18 | 23 | 24 | 45 (59) |
| UPDRS IV score | 12 | 0 | NA | 0 | 0 (59) |
| Hoehn & Yahr score | NA | 2 | 1 | NA | 2 (59) |
| Hoehn & Yahr score ON | 2 | 2 | 1 | 1 | 2 (59) |
| LEDD | 1650.4 | 310 | 0 | 890 | 709.5 |
| **Diagnostics** |  |  |  |  |  |
| DaTSCAN - mean caudate | 1.3399 (54) | 1.395 (61) | 1.815 (40) | 1.705 (55) | 1.1 (52) |
| DaTSCAN - mean putamen | 0.445 (54) | 0.435 (61) | 0.825 (40) | 0.85 (55) | 0.445 (52) |
| DaTSCAN - mean striatum | 0.8925 (54) | 0.915 (61) | 1.3199 (40) | 1.2775 (55) | 0.7725 (52) |
| SAA (CSF) | + | + | + | + | NA |
| Other genetic findings* | *GBA1* p.Leu483Pro | - | - | - | - |
| *APOE* haplotype | E2/E3 | E2/E3 | E3/E3 | E3/E3 | E3/E3 |

\* Other genetic findings only include known variants in *GBA1*, *LRRK2*, and *SNCA*.

#### Individual 1

This male had an age at motor symptom onset (AAO) in his late 40s. Initial motor symptoms included mild unilateral bradykinesia and rigidity (UPDRS III 16 points, H&Y 1; untreated). He also reported mild non-motor signs, including constipation, light-headedness, fatigue, pain, depression, and sleep impairment (UPDRS I 8 points). Levodopa was initiated 3 years after onset; the levodopa-equivalent daily dose (LEDD) at his latest follow-up after 13 years of disease was 1650 mg. Over the disease course, symptoms progressed to bilateral motor involvement, gait impairment with slight freezing, postural instability (UPDRS III 43 points, H&Y 2; ON stage), and motor complications, including dyskinesias and painful off-dystonia. He had mild cognitive impairment (MCI; MOCA 24/30 points in his 60s) and hyposmia. The α-synuclein seed amplification assay (SAA) was positive, and the DaTSCAN showed reduced putaminal uptake with relative caudate sparing, consistent with neurodegenerative PD. Notably, this individual had a positive family history of PD, and he also carried the *GBA1* p.Leu483Pro variant.

#### Individual 2

This individual is a female with PD onset at age of his 50s. Her initial motor symptoms were mild unilateral bradykinesia, rigidity, and tremor (UPDRS III 17 points, H&Y 1; untreated). For non-motor signs, she reported mild constipation and urinary problems (UPDRS I 3 points). She also has hyposmia. She was started on levodopa in the first year after onset, but only required small dosages (LEDD 310 mg at her latest assessment after 7 years of disease duration). Over the course of the disease, she developed slight bilateral motor involvement, but her symptoms remained fairly stable otherwise (UPDRS III 18 points, H&Y 2; ON stage). SAA was positive, and the DaTSCAN showed reduced putaminal uptake with relative caudate sparing, consistent with neurodegenerative PD.

#### Individual 3

The male patient developed first PD symptoms at his 30s. Initial motor symptoms were mild unilateral bradykinesia, rigidity, and tremor (UPDRS III 14 points, H&Y 1; untreated). Non-motor signs were mild and included constipation and sleep impairment (UPDRS I 4 points). He remained untreated over a disease course of 2 years, while his motor symptoms progressed but remained unilateral (UPDRS III 23 points, H&Y 2; ON stage). He had no hyposmia. SAA was positive, and the DaTSCAN showed findings suggestive of early dopaminergic degeneration.

#### Individual 4

This female patient had PD onset at her 50s. Her initial motor symptoms were mild unilateral bradykinesia and rigidity (UPDRS III 15 points, H&Y 1; untreated). Levodopa was initiated in the first year after onset, with a moderate LEDD of 890 mg at her latest assessment after 10 years of disease. Over the course of the disease, she only showed mild motor progression (UPDRS III 24 points, H&Y 1; ON stage) but reported several non-motor signs, including anxiety and depressive episodes, fatigue, pain, and constipation. She also had hyposmia. SAA was positive, and the DaTSCAN showed mildly reduced putaminal uptake with preserved caudate, potentially indicating early or mild dopaminergic dysfunction.

#### Individual 5

The male patient had an AAO in his 40s. Initial symptoms included mild unilateral bradykinesia, rigidity, and tremor (UPDRS III 12 points, H&Y 1; untreated); no non-motor signs except hyposmia (UPDRS I 0 points). Levodopa was initiated 3 years after onset; the LEDD at his last follow-up after a 12-years disease course was 709 mg. Over 12 years of follow-up, he had a progressive disease course with bilateral motor involvement without relevant motor complications (UPDRS III 36 points, H&Y 2; ON stage). The DaTSCAN showed reduced putaminal uptake with relative caudate sparing, consistent with neurodegenerative PD; SAA was not available.

## Discussion

In this study, we report the first systematic screening of intronic *FGF14* GAA repeat expansions in PD, leveraging long-read WGS data from 463 PD patients and 1,627 controls from PPMI, the All of Us Research Program, the CARD Initiative, and the 1000 Genomes Project. We identified five PD patients and one control carrying pathogenic length *FGF14* GAA expansions. A similar frequency in controls was previously reported by Mohren et al., who identified two out of 802 controls carrying pathogenic *FGF14* GAA expansions, supporting the notion of incomplete penetrance within this repeat size range^19^. We also assessed alleles within the reduced penetrance range (250–300 repeats) and characterized alternative, non-pathogenic repeat configurations, including nonpathogenic (GAAGGA), (GAACGA), and composite motifs such as (GAA) (CGA), providing what is, to our knowledge, the most comprehensive population-level analysis to date.

The clinical profiles of the five individuals carrying pathogenic *FGF14* repeat expansions were consistent with PD without pronounced atypical features. While most individuals had a rather mild to moderate disease course, one individual showed marked progression with motor complications and development of MCI. Notably, this individual also carried the *GBA1* p.Leu483Pro variant in addition to a pathogenic *FGF14* repeat expansion. This individual also was the only one with a positive family history of PD. Interestingly, four individuals had an early AAO ≤50 years, with AAO overall ranging from 30s to 50s years; however, there was no correlation between *FGF14* repeat length and AAO. Variable non-motor features were reported, but were generally mild; hyposmia was reported in four individuals. SAA was positive for all four tested individuals, and DaTSCAN imaging consistently demonstrated findings suggestive of neurodegenerative PD. Notably, the clinical assessment was tailored to PD, and possible (mild) cerebellar signs, typical for *FGF14* repeat expansion carriers, may not have been adequately assessed. While one individual carried a *GBA1* variant, none of the identified *FGF14* repeat expansion carriers harbored a disease-explaining genetic variant in a gene linked to monogenic PD.

Our findings should be interpreted in the context of several limitations. First, the current analysis was restricted to individuals of European ancestry. Future studies in diverse populations are needed to better define the global frequency and phenotypic impact of *FGF14* repeat expansions in PD. Second, we observed that the *FGF14* locus is hypermethylated in blood-derived DNA across both carriers and non-carriers, with no visible methylation differences between groups. In contrast, analysis of brain-derived DNA from the cerebellum of the NABEC carrier showed hypomethylation at the *FGF14* locus, whereas no methylation difference was observed between the expanded and non-expanded allele (Supplementary Figure 4). These findings are consistent with existing literature showing that *FGF14* is expressed at low levels in blood and more robustly in the brain. However, interpretation is limited by the availability of only a single brain-derived sample from a confirmed expansion carrier, underscoring the need for additional studies across multiple individuals and brain regions to clarify the regulatory consequences of the repeat expansion. Third, although a pathogenic threshold of ≥300 GAA repeats has been proposed based on prior work in ataxia, the penetrance, expressivity, and mechanistic impact of these expansions in the context of PD remain to be fully defined.

Taken together, our results suggest that pathogenic *FGF14* GAA repeat expansions are present among PD patients of European descent, with an estimated frequency of 1.22 %. While we identified five carriers of fully penetrant *FGF14* repeat expansions, the clinical significance in the context of PD remains unknown. Follow-up analyses, including segregation and functional studies, are required to determine whether this genetic variation plays a causal role in PD or represents a coincidental finding without disease relevance. Integrative analyses incorporating long-read sequencing, transcriptomics, and methylation profiling from brain tissue will be essential to determine the functional relevance of *FGF14* expansions in PD and other neurodegenerative disorders.

## Data availability

Extracted DNA for 1000 Genomes Project was obtained from the Coriell Institute for Medical Research and was consented for the full public release of genomic data. Please see Coriell (https://www.coriell.org) for more information on specific cell lines. 1000 Genomes Project ONT dataset was generated at the Institute of Molecular Pathology (Vienna, Austria) with funds provided by Boehringer-Ingelheim. All of Us genomic data are publicly available to registered researchers on the All of Us Researcher Workbench at https://www.researchallofus.org/data-tools/workbench/. Researchers can apply for access to the All of Us database following the instructions at https://www.researchallofus.org/register/. PPMI data used in the preparation of this article were obtained on 2025-06-01 from the PPMI database (www.ppmi-info.org/access-data-specimens/download-data), RRID: SCR_006431. For up-to-date information on the study, visit www.ppmi-info.org. The PPMI ONT data will be available at the LONI IDA.

## Code availability

The pipeline and analyses presented in this manuscript are publicly available at https://github.com/NIH-CARD/CARDlongread_FGF14_repeat_expansion.

## Acknowledgements

We would like to thank all participants who donated their time and biological samples to this study. We also thank the team at the Parkinson’s Progression Markers Initiative (PPMI) for providing frozen blood samples for long-read DNA sequencing, including Tatiana M. Foroud, Jan E. Hamer, Caitlin D. Schulz, Bradford Casey, and Mark Frasier. The 1000 Genomes Project Consortium ONT panel data were generated at the Institute of Molecular Pathology (Vienna, Austria), with funding from Boehringer-Ingelheim. The authors gratefully acknowledge the All of Us Research Program participants, without whom this research would not be possible.

## Funding

This work was supported in part by the Intramural Research Programs of the National Institute on Aging (NIA) and the National Institute of Neurological Disorders and Stroke (NINDS), National Institutes of Health, Department of Health and Human Services (project numbers Z01-AG000949 and 1ZIANS003154). Computational analyses were performed using the NIH HPC Biowulf cluster (http://hpc.nih.gov).

This research was supported in part by the Intramural Research Program of the National Institutes of Health (NIH). The contributions of the NIH authors were made as part of their official duties as NIH federal employees, are in compliance with agency policy requirements, and are considered Works of the United States Government. However, the findings and conclusions presented in this paper are those of the authors and do not necessarily reflect the views of the NIH or the U.S. Department of Health and Human Services.

Clinical data and biosamples used in this study were obtained from the MJFF Parkinson’s Progression Markers Initiative (PPMI). PPMI – a public-private partnership – is funded by the Michael J. Fox Foundation for Parkinson’s Research and funding partners, including 4D Pharma, Abbvie, AcureX, Allergan, Amathus Therapeutics, Aligning Science Across Parkinson’s, AskBio, Avid Radiopharmaceuticals, BIAL, BioArctic, Biogen, Biohaven, BioLegend, BlueRock Therapeutics, Bristol-Myers Squibb, Calico Labs, Capsida Biotherapeutics, Celgene, Cerevel Therapeutics, Coave Therapeutics, DaCapo Brainscience, Denali, Edmond J. Safra Foundation, Eli Lilly, Gain Therapeutics, GE HealthCare, Genentech, GSK, Golub Capital, Handl Therapeutics, Insitro, Jazz Pharmaceuticals, Johnson & Johnson Innovative Medicine, Lundbeck, Merck, Meso Scale Discovery, Mission Therapeutics, Neurocrine Biosciences, Neuron23, Neuropore, Pfizer, Piramal, Prevail Therapeutics, Roche, Sanofi, Servier, Sun Pharma Advanced Research Company, Takeda, Teva, UCB, Vanqua Bio, Verily, Voyager Therapeutics, the Weston Family Foundation and Yumanity Therapeutics. The PPMI Investigators did not participate in the analysis or preparation of this manuscript. For additional study information, see www.ppmi-info.org.

The All of Us Research Program is supported by the National Institutes of Health, Office of the Director, under multiple cooperative agreements (see full list below*).

K.D. was supported in part by the JSPS Research Fellowship for Japanese Biomedical and Behavioral Researchers at NIH. K.J.B. was supported in part by the William H. Gates Sr. Fellowship from the Alzheimer’s Disease Data Initiative.

*All of Us funding acknowledgments:

Regional Medical Centers: 1 OT2 OD026549; 1 OT2 OD026554; 1 OT2 OD026557; 1 OT2 OD026556; 1 OT2 OD026550; 1 OT2 OD026552; 1 OT2 OD026553; 1 OT2 OD026548; 1 OT2 OD026551; 1 OT2 OD026555; IAA #: AOD 16037;

Federally Qualified Health Centers: HHSN 263201600085U;

Data and Research Center: 5 U2C OD023196;

Biobank: 1 U24 OD023121;

The Participant Center: U24 OD023176;

Participant Technology Systems Center: 1 U24 OD023163;

Communications and Engagement: 3 OT2 OD023205; 3 OT2 OD023206;

Community Partners: 1 OT2 OD025277; 3 OT2 OD025315; 1 OT2 OD025337; 1 OT2 OD025276.

---

**Supplementary Figure 1.**
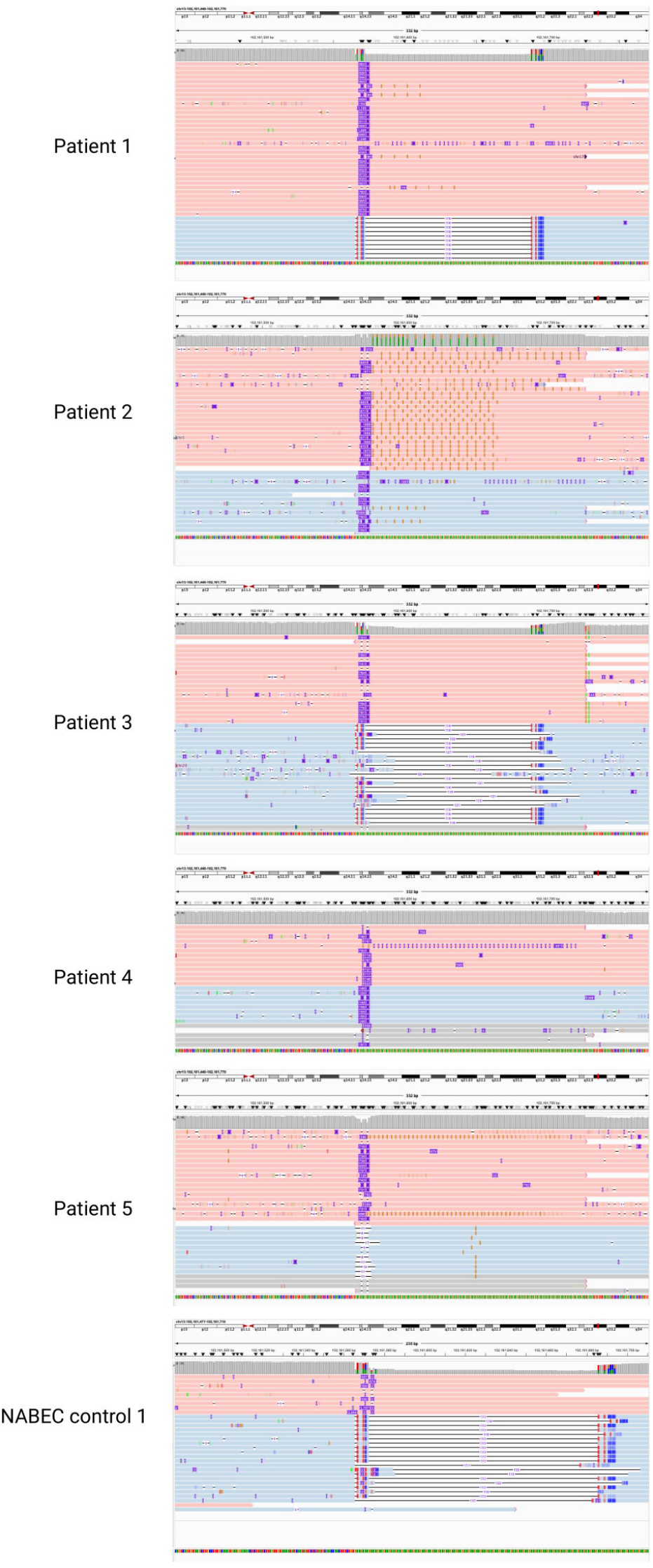
Integrative Genomics Viewer visualization of the FGF14 (GAA) repeat expansion in affected carriers. Genome browser snapshots showing aligned long-read sequencing reads at the *FGF14* locus for the five PD patients carrying the pathogenic expansion. Expanded alleles are indicated by increased repeat length relative to the reference.

**Supplementary Figure 2.**
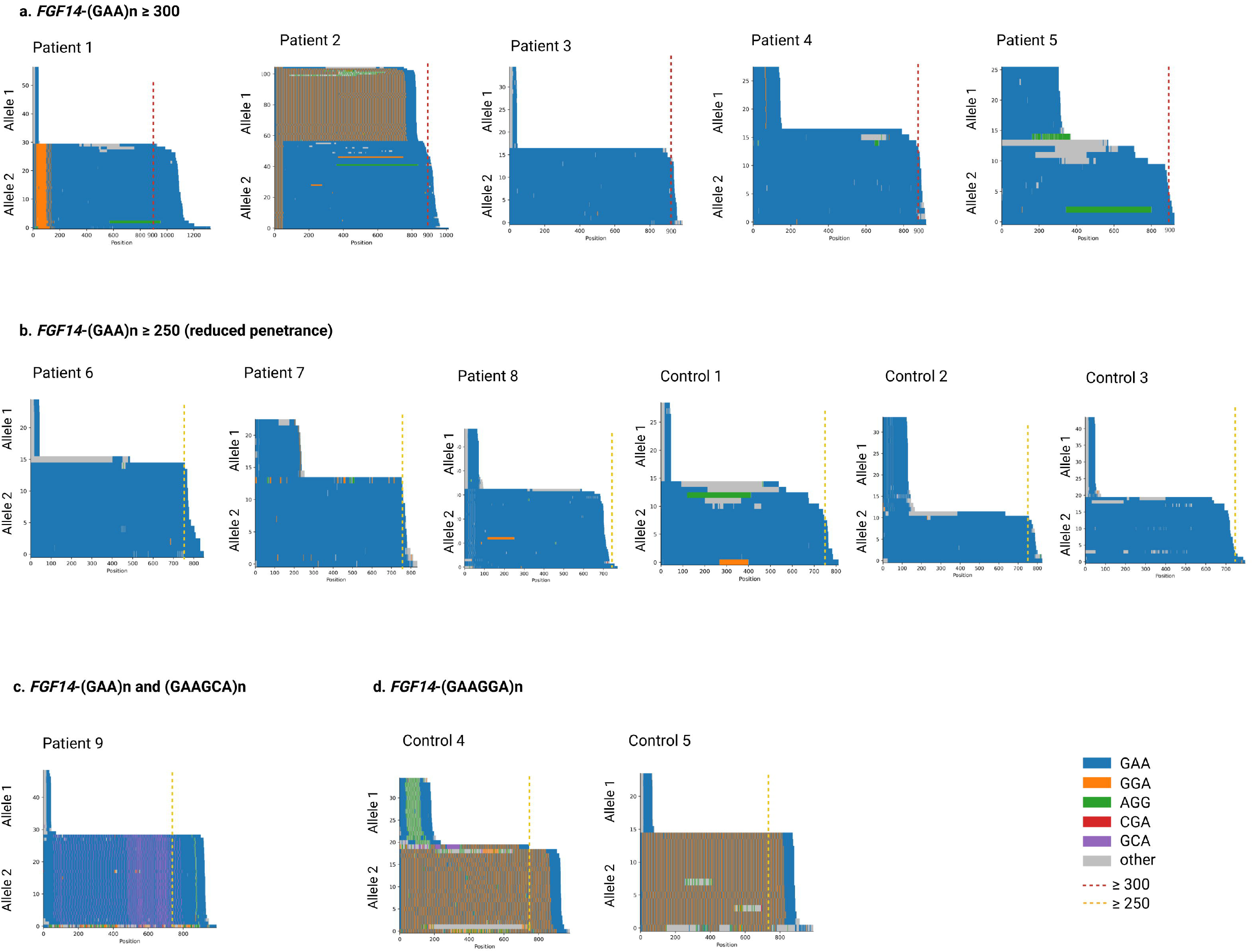
Pathogenic and reduced penetrance *FGF14* (GAA)lll repeat expansions identified in PPMI Parkinson’s disease patients. **a)** Waterfall plot displaying the repeat lengths observed in five Parkinson’s disease cases carrying fully penetrant (GAA) expansion ≥300 repeat units, **b)** three patients and three healthy controls with reduced penetrance *FGF14* (GAA) ≥ 250 repeat units, **c)** one patient with (GAAGCA) motif, **d)** two controls with (GAAGGA) expansions.

**Supplementary Figure 3.**
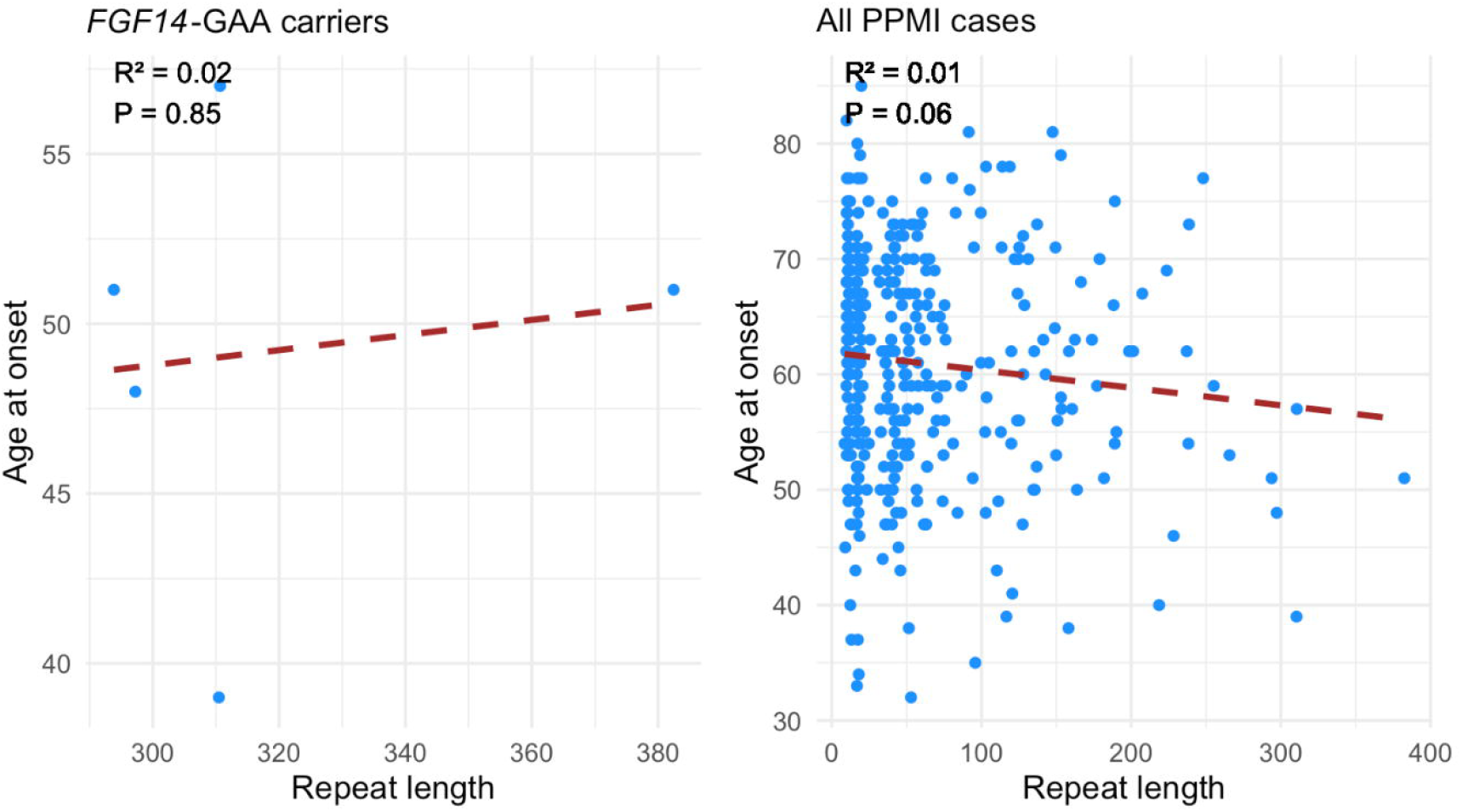
Distribution of age at onset by *FGF14* (GAA)lll repeat length. Analysis of five *FGF14*-GAA expansion carriers from the PPMI cohort (left), and analysis of all PPMI Parkinson’s disease cases with available age at onset and repeat length data (right).

**Supplementary Figure 4.**
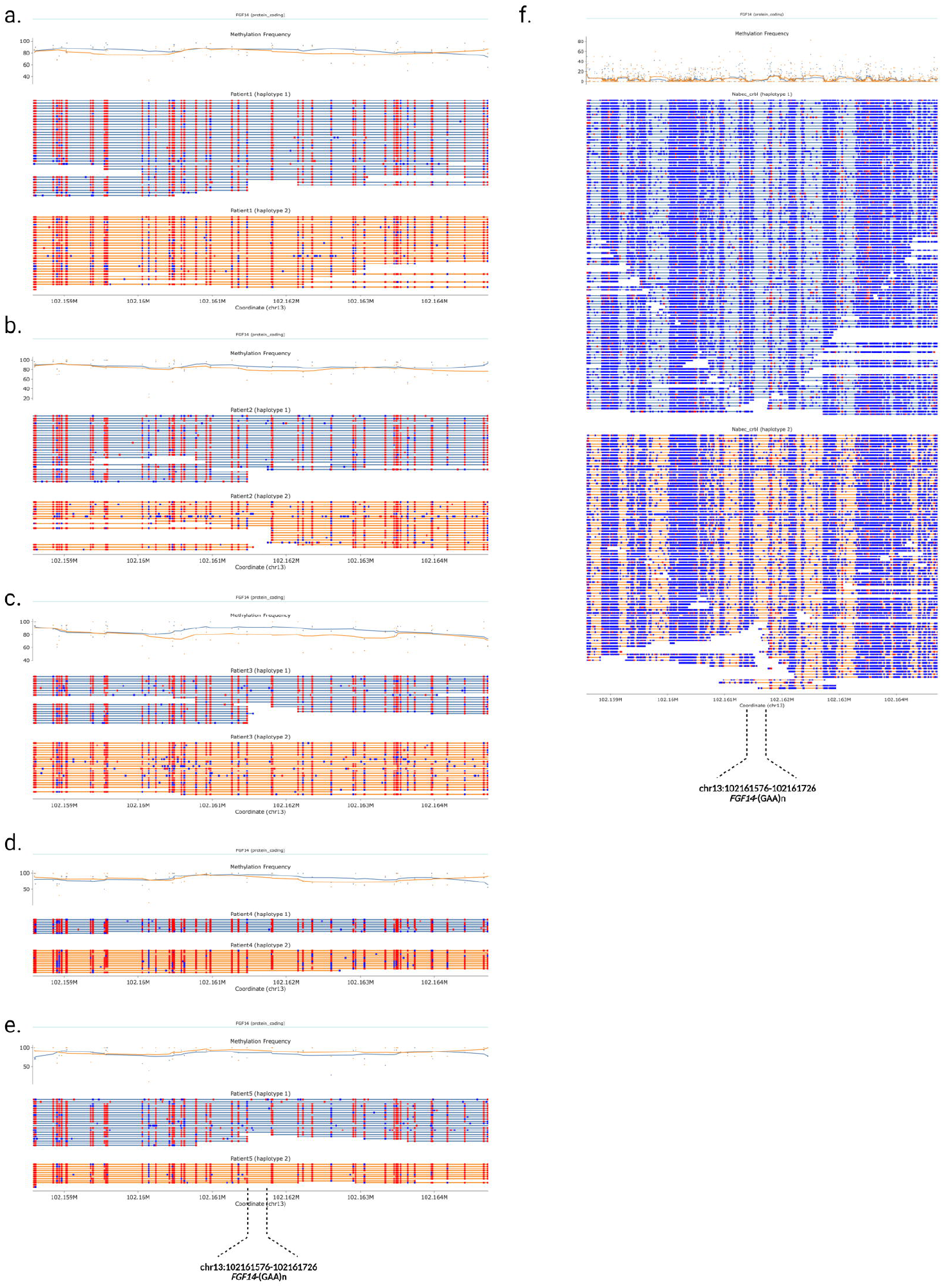
Haplotype-specific DNA methylation across the *FGF14* (GAA)lll expansion a–e) Methylation frequency plots generated with modbamtools for PD patients heterozygous for a pathogenic *FGF14* (GAA) expansion, based on PPMI blood-derived long-read sequencing data. **f)** Methylation frequency plot for the control individual from the NABEC cohort, based on adaptive sampling from cerebellum tissue. Haplotypes are phased, with haplotype 1 corresponding to the non-expanded allele and haplotype 2 representing the expanded allele. Methylation frequency is shown above, with the *FGF14* gene structure overlaid. Individual reads are shown below, with blue indicating hypomethylation and red indicating hypermethylation.

**Supplementary Table 1.**
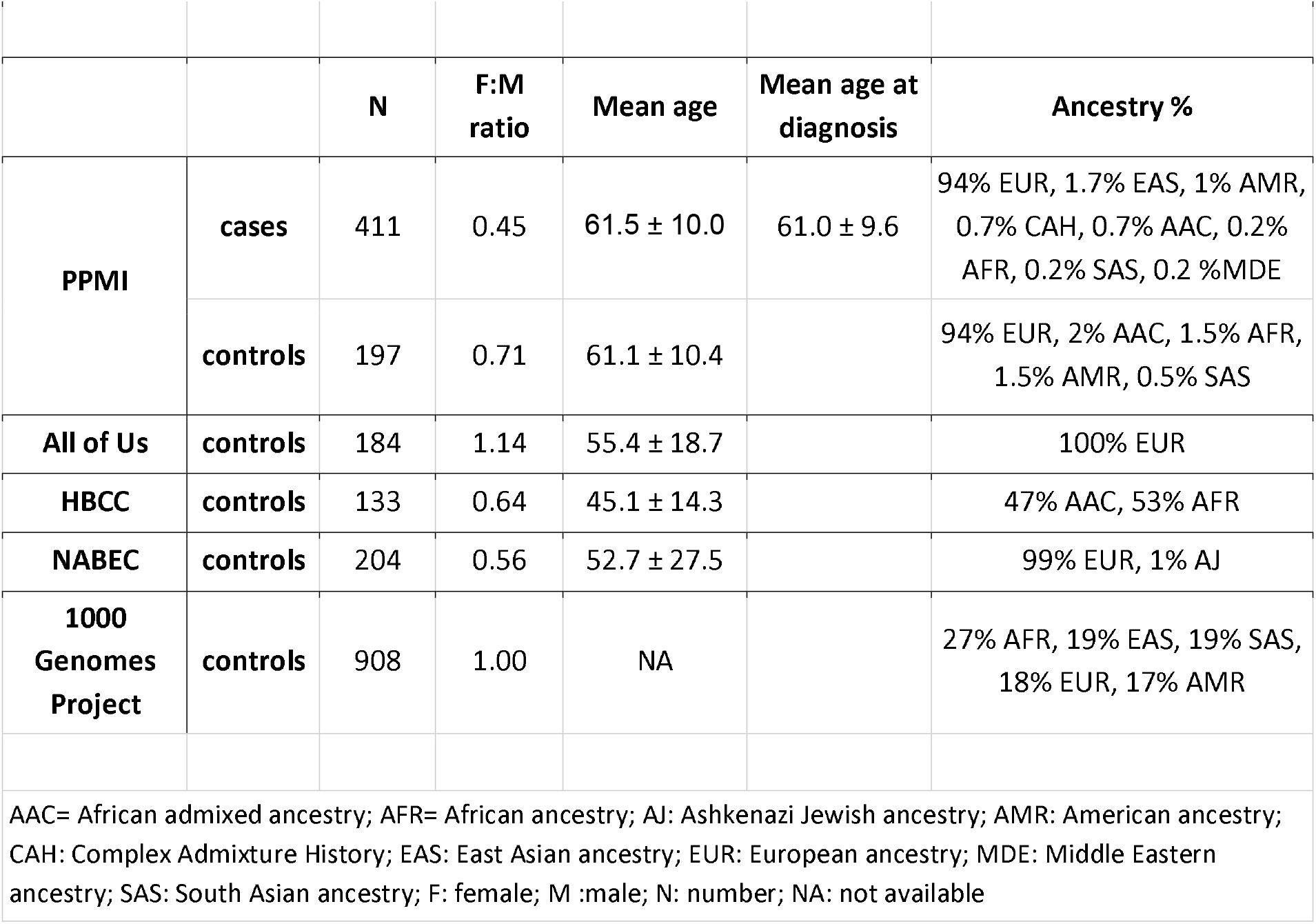
Demographic characteristics of study participants (PPMI, All of Us, HBCC, NABEC, 1000 Genomes Project).

**Supplementary Table 2. Summary of *FGF14* repeat expansion in 411 PPMI cases and 1,626 controls from PPMI, NABEC, HBCC, All of Us, and 1,000 Genomes Project participants**. Length is defined by the average of the ten longest alleles carrying the expansions.

