## Supplementary Methods for "Long-read sequencing identifies *FGF14* repeat expansions in Parkinson’s disease"

**DNA Extraction and Library Preparation**

High molecular weight (HMW) DNA was extracted from 1 mL of frozen whole blood using the KingFisher Apex instrument and an adapted version of the PacBio Nanobind HT protocol for HMW DNA extraction (see detailed protocol: [dx.doi.org/10.17504/protocols.io.x54v9py8qg3e/v1](https://dx.doi.org/10.17504/protocols.io.x54v9py8qg3e/v1)). The Nanobind HT 1 mL Blood Kit (PacBio, 102-762-800) was used. To enrich long DNA fragments, a size-selection step was performed using the PacBio Short Read Eliminator Kit (102-208-300) to remove fragments smaller than 25 kb.

The DNA was then sheared to a target length of 30 kb using the Megaruptor 3 instrument (Diagenode) with the standard Megaruptor 3 Shearing Kit (E07010003). Libraries were prepared using the Oxford Nanopore Technologies (ONT) SQK-LSK114 Ligation Sequencing Kit and sequenced on PromethION R10 flow cells (FLO-PRO114M) for 72 hours following ONT standard operating procedures.

**Data Generation and Processing**

Raw signal data (FAST5 or POD5 files) were generated using MinKNOW v22.10.7. Basecalling was performed using Dorado v0.7.1 (ONT) with the dna_r10.4.1_e8.2_400bps_sup@v5.0.0 model, which includes detection of 5mCG and 5hmCG methylation. Reads passing quality control during basecalling were aligned to the GRCh38 (hg38) reference genome using minimap2 with the map-ont preset. Resulting SAM files were converted to BAM format, sorted, and indexed with Samtools v1.11.

To identify potential sample swaps, SNVs were called per flow cell using Clair3 v1.0.4. Variant calls were merged using bcftools, and kinship coefficients were calculated using PLINK2. Flow cell runs with the same sample name, a kinship coefficient >0.375, and >75,000 shared SNVs were flagged as potential swaps. These were resolved prior to downstream analysis.

After resolving swaps, unmapped BAMs per sample were merged, sorted, and indexed using samtools to generate a single BAM file per sample. Alignments with an average Q score <10 were removed using samtools view.

**Validation of expansion carriers with adaptive sampling**

To validate and more precisely characterize *FGF14* repeat expansions, we performed further targeted ONT adaptive sampling on genomic DNA from the five individuals in the PPMI cohort and one control individual from the NABEC cohort. High molecular weight DNA was extracted from blood for the five PPMI patients and from cerebellum tissue for the NABEC control. Libraries were prepared using the ONT ligation sequencing kit (SQK-LSK114). Adaptive sampling was performed on PromethION R10 flow cells using a custom BED file targeting the *FGF14* locus (chr13:94060103-110056532, hg38) to enrich for reads spanning the repeat region. Basecalling was carried out with dorado v1.0.0 using the super accuracy model and 5mC/5hmC modification calling (dna_r10.4.1_e8.2_400bps_sup@v5.0.0, 5mC_5hmC), excluding reads with an average basecalling Q score of under 10. Reads were aligned to the GRCh38 reference genome using minimap2 (v2.26), and repeat sizes were re-estimated using Straglr. Repeat motif structure and interruptions were visualized using RepeatAnalysisTools.

**Methylation analysis**

To assess DNA methylation patterns at the *FGF14* locus, we employed modbamtools (v0.4.8)^20^ to visualize haplotype-resolved methylation profiles in repeat expansion carriers and a control individual. Haplotype-tagged BAM files generated by the PEPPER-Margin-DeepVariant pipeline (https://github.com/kishwarshafin/pepper) were used as input, alongside gene annotation from Gencode v38 (GRCh38). The --hap flag was included to enable separation of methylation frequencies by haplotype, allowing comparison between expanded and non-expanded alleles.
